# The immune checkpoint pathophysiology of depression and chronic fatigue syndrome due to preeclampsia: focus on sCD80 and sCTLA-4

**DOI:** 10.1101/2024.04.14.24305809

**Authors:** Jangir Sami Omar, Niaz Albarzinji, Mengqi Niu, Naz Hawree Taher, Bayar Aram, Mohammed Salam Sulaiman, Shatha Rouf Moustafa, Hussein Kadhem Al-Hakeim, Michael Maes

**Affiliations:** College of Medicine, Hawler Medical University, Erbil, Iraq; Erbil Center KHCMS, College of Medicine, Hawler Medical University, Erbil, Iraq; Sichuan Provincial Center for Mental Health, Sichuan Provincial People’s Hospital, School of Medicine, University of Electronic Science and Technology of China, Chengdu 610072, China; Key Laboratory of Psychosomatic Medicine, Chinese Academy of Medical Sciences, Chengdu, 610072, China; College of Pharmacy, Hawler Medical University, Erbil, Iraq; Baharka Hospital, Erbil, Iraq; Hawler Medical University, College of Pharmacy, Erbil, Iraq; Clinical Analysis Department, College of Pharmacy, Hawler Medical University, Havalan City, Erbil, Iraq; Department of Chemistry, College of Science, University of Kufa, Iraq; Department of Psychiatry, Faculty of Medicine, Chulalongkorn University, Bangkok, Thailand; Department of Psychiatry, Medical University of Plovdiv, Plovdiv, Bulgaria; Research Institute, Medical University of Plovdiv, Plovdiv, Bulgaria; Kyung Hee University, Seoul, Dongdaemun-gu, South Korea

**Keywords:** Neuroimmune, inflammation, chronic fatigue syndrome, affective disorders, biomarkers, pathways

## Abstract

**Background:** Neuropsychiatric disorders in preeclampsia (PE) women are prevalent and worsen PE outcome. Immune-related biomarkers including soluble sCD80 and cytotoxic T-lymphocyte antigen-4 (sCTLA-4) are not well studied in relation to depression, anxiety, and chronic fatigue due to PE.

**Aims:** To study serum immune-inflammatory biomarkers of PE and delineate their associations with the Hamilton Depression (HAMD), Anxiety (HAMA), and Fibro-Fatigue (FF) rating Scale scores.

**Methods:** sCD80, sCTLA-4, vitamin D, granulocyte-macrophage colony-stimulating factor, zinc, copper, magnesium, and calcium were measured in 90 PE compared with 60 non-PE pregnant women.

**Results:** PE women show higher depression, anxiety and FF rating scale scores as compared with control women. sCTLA-4, sCD80, and copper were significantly higher and zinc, magnesium, and calcium significantly lower in PE women than in controls. Multiple regression analysis showed that around 55.8%-58.0% of the variance in the HAMD, HAMA and FF scores was explained by the regression on biomarkers; the top 3 most important biomarkers were sCTLA-4, sCD80, and vitamin D. The sCTLA-4/sCD80 ratio was significantly and inversely associated with the HAMD/HAMA/FF scores. We found that around 70% of the variance in systolic blood pressure could be explained by sCTLA-4, vitamin D, calcium, and copper.

**Conclusions:** The findings underscore that PE and depression, anxiety, and chronic fatigue symptoms due to PE are accompanied by activation of the immune-inflammatory response system. More specifically, disbalances among soluble checkpoint molecules seem to be involved in the pathophysiology of hypertension and neuropsychiatric symptoms due to PE.

## Introduction

Preeclampsia (PE) is a common and potentially fatal condition that manifests during pregnancy. It is distinguished by the abrupt onset of hypertension, cephalalgia, and visual impairments (1–3). Fifteen percent of annual maternal fatalities in developing countries are attributed to PE, according to estimates (4). Reportedly, in addition to pain, hypertension, and edema, women with PE experience chronic fatigue, depression, and anxiety (5–10). Furthermore, several research studies have demonstrated a correlation between the severity of PE symptoms and an elevated prevalence of depression (5, 11).

PE is characterized by endothelial dysfunctions, immune abnormalities, and syncytiotrophoblast stress (12). PE is frequently associated with biomarkers of oxidative stress, inflammation, immune activation, and autoimmune responses (13, 14). Placental apoptosis and necrosis may result from chronic hypoxia in the intervillous region, which may induce oxidative stress in the tissues (15). Pro-inflammatory T helper (Th)1 and Th17 cytokines, along with suppressive Treg and Th2 cytokines (IL-10 and IL-4), have been found to be associated with PE at both the systemic and local levels (16–18). The transition to a Th1 response, characterized by increased IFN-γ secretion, is a critical element in PE (19).

The cytotoxic T-lymphocyte antigen-4 (CTLA-4 or CD152) gene may serve as a risk factor for PE during pregnancy, according to the findings of prior research (20). CTLA-4 is a protein receptor that downregulates immunological responses and functions as an immune checkpoint (21). An inhibitory signal is generated when CLTA-4 binds to cluster of differentiation 80 (CD80) (or CD86), which is expressed on antigen-presenting cells. This signal prevents the activation of CD28 (22). CD80, classified as a B7, type I membrane protein within the immunoglobulin superfamily, functions as a costimulatory molecule for T-cells and is implicated in T-cell activation (23, 24). Interestingly, soluble CTLA-4 (sCTLA-4) and CD80 (sCD80) are measurable in serum (25, 26), and have roles in modulating the immune response and autoimmune responses (27–30). High concentrations of sCTLA-4 were observed in sera of patients with autoimmune thyroid diseases (31), as well as in patients with type 1 diabetes, diffuse cutaneous systemic sclerosis (32), systemic lupus erythematosus (33), and rheumatic arthritis (34). An increase in sCD80 levels may lead to an increase in IFN-γ production by active T cells (35). sCD80 levels increased significantly in patients with autoimmune disease including SLE (36) and rheumatoid arthritis (37) compared to the healthy population.

Granulocyte-macrophage colony-stimulating factor (GM-CSF) is an additional immune biomarker that should be considered in the context of PE. Endothelial cells, macrophages, mast cells, T cells, and fibroblasts are responsible for producing the latter glycoprotein (38–40). The induction of differentiation and activation of macrophage and dendritic cells by GM-CSF suggests that it might play a crucial role in the pathogenesis of PE (41). GM-CSF, lipopolysaccharide (LPS) and/or IFN-γ polarize macrophages toward an M1 phenotype, which is characterized by the increased expression of proinflammatory cytokines and CD80 (42, 43). Prevalence estimates for vitamin D deficiency during pregnancy range from 8 to 70%, contingent upon factors such as UV exposure and skin pigmentation (44, 45). Vitamin D deficiency is more prevalent among mothers with PE and their neonates; therefore, patients may be advised to take higher doses of vitamin D supplementation (46, 47).

An activated immune-inflammatory response (IRS), which includes Th1 and Th17 responses, has been found to be associated with affective disorders and chronic fatigue syndrome (CFS) (48–50). Reduced levels of albumin, zinc, calcium, and magnesium accompany this IRS response (51, 52). Furthermore, it has been observed that prenatal chronic fatigue and perinatal depression are associated with decreased serum zinc and IRS activation (53, 54). However, the correlations between affective symptoms and CFS due to PE and immune biomarkers, including sCTLA4, sCD80, vitamin D, zinc, copper, albumin, calcium, and magnesium, remain largely unknown.

Therefore, the current study aimed to examine the correlations between immune-related biomarkers (sCD80, sCTLA-4, GM-CSF, vitamin D, zinc, calcium, magnesium, copper) in women with PE versus controls and their associations with depression, anxiety, and CFS due to PE.

## Subjects and Methods

### Subjects

From November 2022 to February 2023, the present study recruited sixty healthy expectant control women of comparable age and gestational age and ninety PE women with an average age of 32.67±5.88 years. The participants were recruited from maternity teaching institutions and selected private clinics. The diagnosis of PE was made in accordance with the criteria established by the American College of Obstetricians and Gynecologists (55). After 20 weeks of pregnancy, PE was identified in women who exhibit proteinuria and have a systolic and diastolic blood pressure higher than 140 mmHg and 90 mmHg, respectively. Each patient in the investigation fulfilled the specified criteria, and proteinuria was detected in all cases using dipstick tests. In addition, the patients were administered methyldopa (Aldomet^®^) and were required to fast overnight. The patient’s gravidity was characterized as the cumulative count of pregnancies, encompassing abortions, ectopic pregnancies, and any other pregnancies recorded in the medical record. Parity denotes the count of births that transpire after the 28th week of gestation, encompassing stillbirths and intrauterine fetal fatalities (IUFD). Sixty women who were at least 20 weeks expectant and lacked any PE symptoms were chosen to comprise the control group. The controls were matched for gestational age to the PE patients. Their blood pressure was normal at <120/80 mmHg.

A comprehensive medical history evaluation was conducted on each participant to exclude any pre-existing systemic conditions that could potentially impact the results, including liver and renal disease, infection, and cardiovascular events. All female subjects who were taking immunosuppressants or had compromised immune systems were precluded from the study. There were no prenatal abnormalities observed in any of the participants. There were no reports of active ailments, including uterine contractions or membrane ruptures. Other exclusion criteria for patients and controls were autoimmune and immune disorders including diabetes mellitus type 1, psoriasis, CFS, lupus erythematosus, arthritis, and inflammatory bowel disease. All subjects with axis 1 neuropsychiatric disorders present before the pregnancy were excluded, such as major depression, bipolar disorder, autism, psycho-organic disorder, and substance use disorders. All subjects showed CRP levels below 6 mg/l (56). Patients who ever had suffered from severe phase 2 (pneumonia) or phase 3 (admission into ICU) COVID-19 were excluded to participate. Women who had suffered from mild COVID-19 infection were allowed to participate if the symptoms had resided at least three months before inclusion in this study.

Under Document No. 103/2022, the University of Hawler’s approval committee in Erbil, Iraq, granted ethical approval for the research endeavor. The authorization verifies that the undertaking conforms to the criteria specified in the “International Guideline for Human Research,” in accordance with the stipulations of the Declaration of Helsinki.

### Clinical assessments

The severity of CFS and fibromyalgia was assessed by a senior psychiatrist using the Fibro-Fatigue scale (57). The level of anxiety was evaluated using the Hamilton Anxiety Rating Scale (HAMA) (58). Hamilton’s Depression Rating Scale (HAMD) (59) was completed by every participant to measure severity of depression. The senior psychiatrist conducted semi-structured interviews to collect sociodemographic and clinical information. When diagnosing tobacco use disorder (TUD), DSM-IV-TR criteria were applied. By dividing weight in kilograms by length in meters, BMI was computed.

### Measurements

Fasting venous blood samples were taken from the participants between 8.00 a.m. and 9.00 a.m. and collected into plain tubes. Samples were aliquoted and stored at −80 °C before assay. After separation, the sera were distributed into three Eppendorf^®^ tubes. Serum albumin, calcium, magnesium, copper, and zinc were measured spectrophotometrically using kits supplied by Spectrum Diagnostics Co. (Cairo, Egypt). The CRP latex slide test (Spinreact^®^, Barcelona, Spain) was used for CRP assays in human serum. The test is based on the principle of latex agglutination. ELISA sandwich kits, supplied by Nanjing Pars Biochem Co., Ltd. (Nanjing, China), were used to measure serum sCD80, GM-CSF, sTCLA-4, and vitamin D. The procedures were followed exactly without modifications according to the manufacturer’s instructions. The intra-assay coefficients of variation (CV) (precision within an assay) were < 10.0%. We computed two indices: a) the ratio of sCTLA-4/sCD80; and b) a z-unit based composite as z sCTLA-4 + z sCD80.

### Statistical analysis

The researchers utilized analysis of variance (ANOVA) to examine the variations in scale variables between control and PE women, while analysis of contingency tables (χ^2^-test) was employed to determine the relationships between nominal variables. To ascertain the impact of diagnosis on the biomarkers, we utilized multivariate general linear model (GLM) analysis, which accounted for confounding variables such as age and BMI. As a result, we conducted between-subjects effect tests in order to examine the associations between the diagnosis and biomarkers. Estimated marginal mean (SE) values generated by the model using GLM analysis were calculated. By utilizing manual and stepwise multiple regression analysis, the biomarkers that best predict the symptoms were identified. Collinearity was assessed in all regression analyses through the utilization of tolerance and VIF values. We employed a manual method and an automatic stepwise approach that incorporated variables with a p-to-entry of 0.05 and a p-to-remove of 0.06. In cases where homoscedasticity was deemed invalid through comprehensive examination of plots comparing standardized residuals to standardized predicted values and the White and Breusch-Pagan test, we employed heteroscedasticity-consistent standard error (SE) or robust SE estimates (utilizing the HC3 method). All analyses were checked using bootstrapped methods (n=1000), and discrepancies between the bootstrapped and other approaches are reported if needed. For statistical significance, two-tailed tests were conducted using a p-value of 0.05. All statistical analyses were conducted utilizing version 29 of IBM SPSS for Windows.

The estimated a priori sample size was calculated using G*Power 3.1.9.4 and applied to the primary analysis, which involved conducting a multiple regression analysis of the rating scale scores on the biomarkers. Based on an effect size of f=0.11 (which accounts for approximately 10% of the variance), along with a maximum of 6 explanatory variables, an alpha value of 0.05, and a power of 0.8, it was determined that a minimum sample size of 130 was necessary. It should be added that the post-hoc estimated power for the same analysis was 1.0.

## Results

### 1. Sociodemographic and clinical data

The results of demographic and clinical data of the healthy controls (HC) and PE patients are presented in **Table 1**. The duration of symptoms in the PE group is 8.0±3.5 weeks and the age of onset at 29.3±5.2 years. BMI, education level, residency, smoking status, number of pregnancies, and gestational age did not significantly differ between PE patients and the control group. PE patients show a significantly increase in systolic and diastolic blood pressure compared with the control group. PE patients have a significantly higher abortion rate, FF score, HAMA score, and HAMD score compared with the non-PE pregnant group. The PE group has a lower number of pregnancies and higher abortion rates than the control group.

**Table 1.** Sociodemographic and clinical parameters in preeclampsia (PE) women and healthy pregnant women groups.

| **Variables** |  | **Control**<br>**(n=60)** | **PE**<br>**(n=90)** | **F/$\chi^2$** | **df** | **p** |
| --- | --- | --- | --- | --- | --- | --- |
| Age | Yrs | 30.7±6.3 | 32.7±5.9 | 3.94 | 1/148 | 0.049 |
| BMI | kg/m<sup>2</sup> | 30.71±3.30 | 31.40±4.29 | 1.12 | 1/148 | 0.291 |
| Smoking | No/Yes | 58/2 | 87/3 | 0 | 1 | 1 |
| Rural/Urban |  | 28/32 | 40/50 | 0.07 | 1 | 0.789 |
| Education | Yrs | 6.10±3.32 | 6.20±3.07 | 0.04 | 1/148 | 0.850 |
| Systolic B.P | mmHg | 117.98±2.27 | 156.88±11.50 | MWU | - | <0.001 |
| Diastolic B.P | mmHg | 78.37±3.16 | 89.89±4.51 | MWU | - | <0.001 |
| Gestational age | Wks | 29.60±4.84 | 30.10±4.39 | 0.43 | 1/148 | 0.513 |
| Duration Symptoms | Wks | 0 | 8.01±3.55 | - | - | - |
| No of pregnancies |  | 2.3±1.4 | 1.8±1.2 | 5.02 | 1/148 | 0.027 |
| Previous abortion | No/Yes | 50/10 | 38/52 | 25.09 | 1 | <0.001 |
| Age of Onset | Yrs. | 0 | 29.3±5.2 | - | - | - |
| FF score |  | 7.7±2.3 | 21.7±5.3 | MWU | - | <0.001 |
| HAMA score |  | 6.0±2.4 | 16.4±4.9 | MWU | - | <0.001 |
| HAMD score |  | 5.0±1.3 | 14.6±4.8 | MWU | - | <0.001 |

MWU: Mann-Whitney U test; BMI: body mass index, B.P: blood pressure, HAMA: Hamilton Anxiety Rating Scale, HAMD: Hamilton Depression Rating Scale, FF: fibro fatigue scale.

### 2. Biomarkers of PE

**Table 2** shows the results of multivariate GLM analysis which examines the associations between the biomarkers (albumin, magnesium, calcium, Vitamin D, sCTLA-4, GM-CSF, sCD80, zinc, and copper) and the diagnosis (PE versus HC) while adjusting for age and BMI. Tests for between-subject effects showed that there were significant associations between diagnosis and (in descending order of importance) vitamin D, sCTLA-4, calcium, sCD80, copper, zinc, albumin, and magnesium.

**Table 2.**
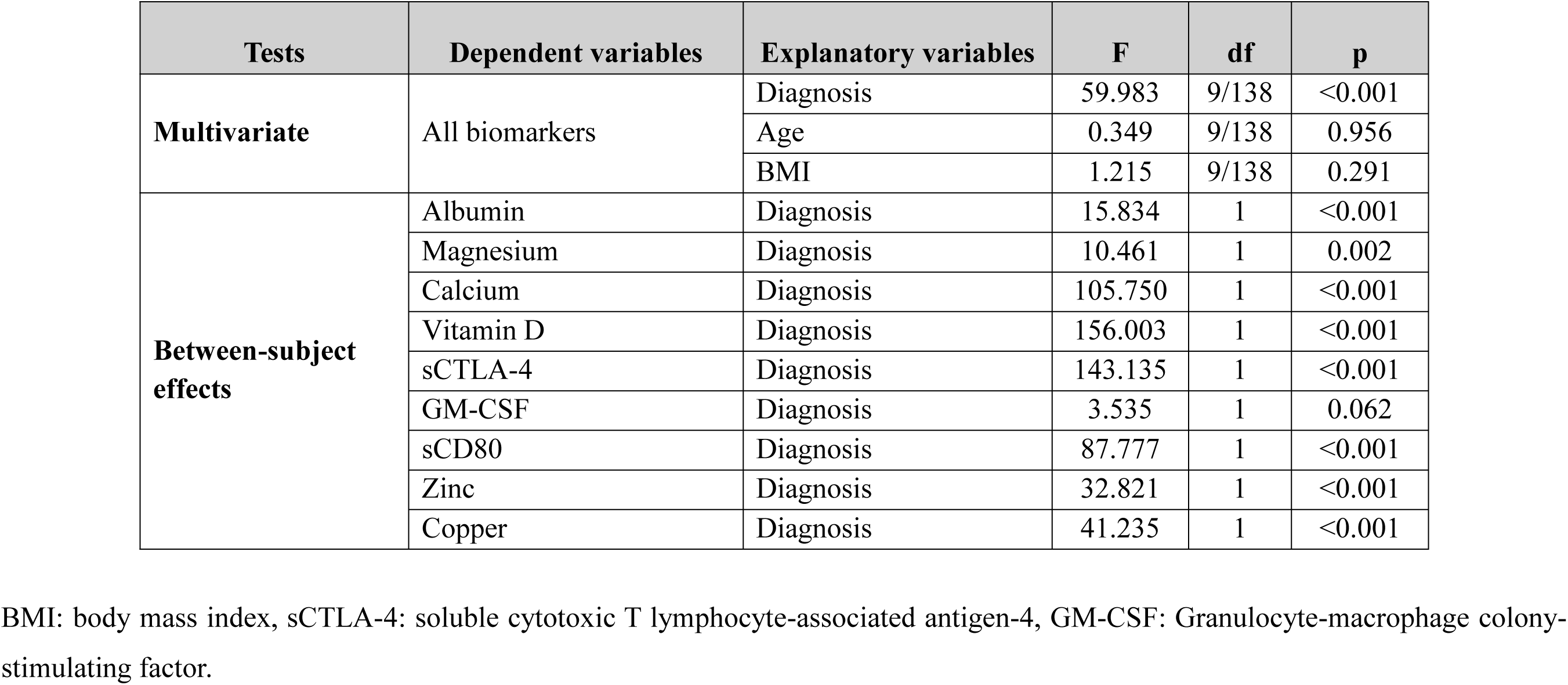
Results of multivariate GLM analysis that examine the associations between the biomarkers and the diagnosis of preeclampsia.

**Table 3** shows the model-generated estimated mean (± SE) biomarker values in the study groups. Serum levels of sCTLA-4, sCD80, copper, and z sCTLA-4 + z sCD80 (z score) show a significant increase in the PE group compared with the control group. There are significant decreases in serum levels of albumin, magnesium, calcium, Vitamin D, zinc, and sCTLA-4 / sCD80 ratio in PE women compared with the healthy pregnant women group.

**Table 3.** Model-derived estimated marginal means of the biomarkers in pre-eclampsia (PE) patients and control pregnant women.

| **Biomarkers** | **Control** | **PE** | **p-value** |
| --- | --- | --- | --- |
| **Albumin** g/l | 48.29(0.77) | 44.32(0.62) | <0.001 |
| **Magnesium** mM | 0.984(0.029) | 0.861(0.024) | 0.002 |
| **Calcium** mM | 2.480(0.021) | 2.201(0.017) | <0.001 |
| **Vitamin D** ng/ml | 12.88(0.30) | 8.02(0.24) | <0.001 |
| **sCTLA-4** pg/ml | 365.9(17.6) | 639.1(14.3) | <0.001 |
| **GM-CSF** pg/ml | 170.5(15.1) | 207.5(12.3) | 0.062 |
| **sCD80** pg/ml | 265.4(19.2) | 4990.(15.6) | <0.001 |
| **Zinc** ppm | 0.775(0.020) | 0.624(0.017) | <0.001 |
| **Copper** ppm | 0.906(0.039) | 1.231(0.032) | <0.001 |
| **sCTLA-4 / sCD80\*** | 1.915±0.140 | 1.419±0.114 | 0.007 |
| **z sCTLA-4 + z sCD80\* (z score)** | -1.622±0.130 | 1.081±0.106 | <0.001 |

\*Results of univariate GLM with age and body mass index as covariates; all other comparisons: results of tests of between-subjects effects after performing multivariate GLM (see Table 2). sCTLA-4: soluble cytotoxic T lymphocyte-associated antigen-4, GM-CSF: granulocyte-macrophage colony-stimulating factor.

### 3. Multiple regression analyses of clinical scores on biomarkers

**Table 4** shows the results of different multiple regression analyses with the psychiatric rating scale scores as dependent variables and blood pressure and biomarkers as explanatory variables, while allowing for the effects of age, BMI, and education. Regression #1 shows that 68.2% of the variance in the total FF score was explained by the regression on systolic and diastolic blood pressure and sCD80 (all positively) and magnesium (inversely). In Regression #2, 62.4% of the variance in the HAMA score was explained by the regression on systolic and diastolic blood pressure, sCD80, and copper (all positively), and albumin (inversely). Regression #3 shows that 59.4% of the variance in the total HAMD score was explained by the regression on systolic and diastolic BP, sCD80, and sCTLA-4.

**Table 4.**
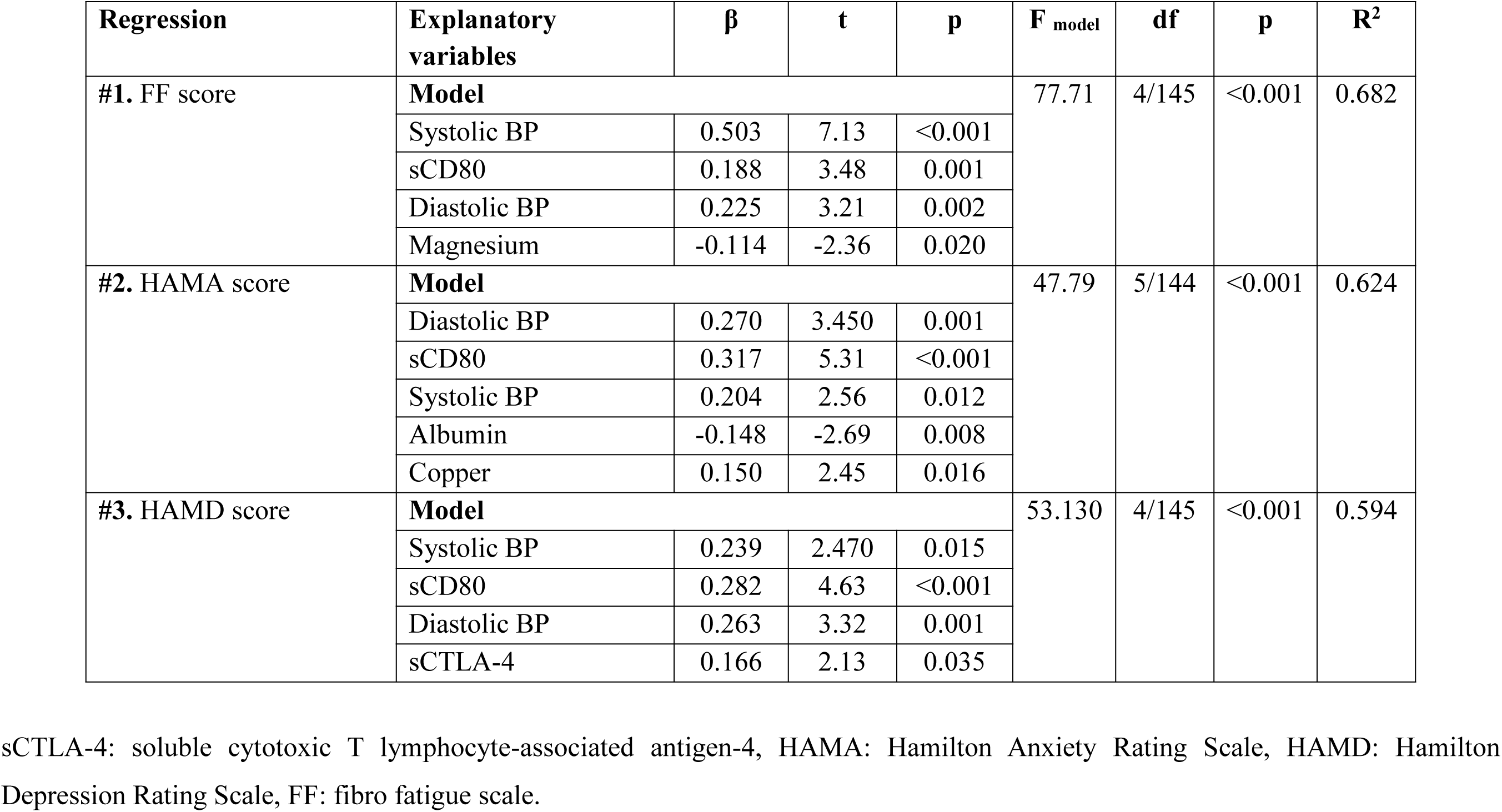
Results of multiple regression analysis with neuropsychiatric rating scale scores as dependent variables and biomarkers and blood pressure (BP) data as explanatory variables.

| **Regression** | **Explanatory variables** | **$\beta$** | **t** | **p** | **F<sub>model</sub>** | **df** | **p** | **R<sup>2</sup>** |
| --- | --- | --- | --- | --- | --- | --- | --- | --- |
| **#1. FF score** | **Model** |  |  |  | 77.71 | 4/145 | <0.001 | 0.682 |
|  | Systolic BP | 0.503 | 7.13 | <0.001 |  |  |  |  |
|  | sCD80 | 0.188 | 3.48 | 0.001 |  |  |  |  |
|  | Diastolic BP | 0.225 | 3.21 | 0.002 |  |  |  |  |
|  | Magnesium | -0.114 | -2.36 | 0.020 |  |  |  |  |
| **#2. HAMA score** | **Model** |  |  |  | 47.79 | 5/144 | <0.001 | 0.624 |
|  | Diastolic BP | 0.270 | 3.450 | 0.001 |  |  |  |  |
|  | sCD80 | 0.317 | 5.31 | <0.001 |  |  |  |  |
|  | Systolic BP | 0.204 | 2.56 | 0.012 |  |  |  |  |
|  | Albumin | -0.148 | -2.69 | 0.008 |  |  |  |  |
|  | Copper | 0.150 | 2.45 | 0.016 |  |  |  |  |
| **#3. HAMD score** | **Model** |  |  |  | 53.130 | 4/145 | <0.001 | 0.594 |
|  | Systolic BP | 0.239 | 2.470 | 0.015 |  |  |  |  |
|  | sCD80 | 0.282 | 4.63 | <0.001 |  |  |  |  |
|  | Diastolic BP | 0.263 | 3.32 | 0.001 |  |  |  |  |
|  | sCTLA-4 | 0.166 | 2.13 | 0.035 |  |  |  |  |

sCTLA-4: soluble cytotoxic T lymphocyte-associated antigen-4, HAMA: Hamilton Anxiety Rating Scale, HAMD: Hamilton Depression Rating Scale, FF: fibro fatigue scale.

In **Table 5**, we have recomputed these associations after deleting the blood pressure data. A significant part of the variance (58.0%) in the total HAMD score can be explained by the regression on sCTLA-4, sCD80, and BMI (all positively), vitamin D, calcium, and GM-CSF (negatively) (regression #1). **Figure 1** shows the partial regression plot of the HAMD total score on serum sCTLA-4. Regression #2 shows that 56.2% of the variance in the HAMA total score was explained by the regression on sCD80, sCTLA-4, copper (all positively), and albumin and vitamin D (both negatively). **Figure 2** shows the partial regression plot of the HAMA total score on serum sCD80. In Regression #3, 55.8 % of the variance in the FF score could be explained by the regression on sCTLA-4, copper, sCD80 (all positively), vitamin D, calcium, and magnesium (inversely associated).

**Figure 1.**
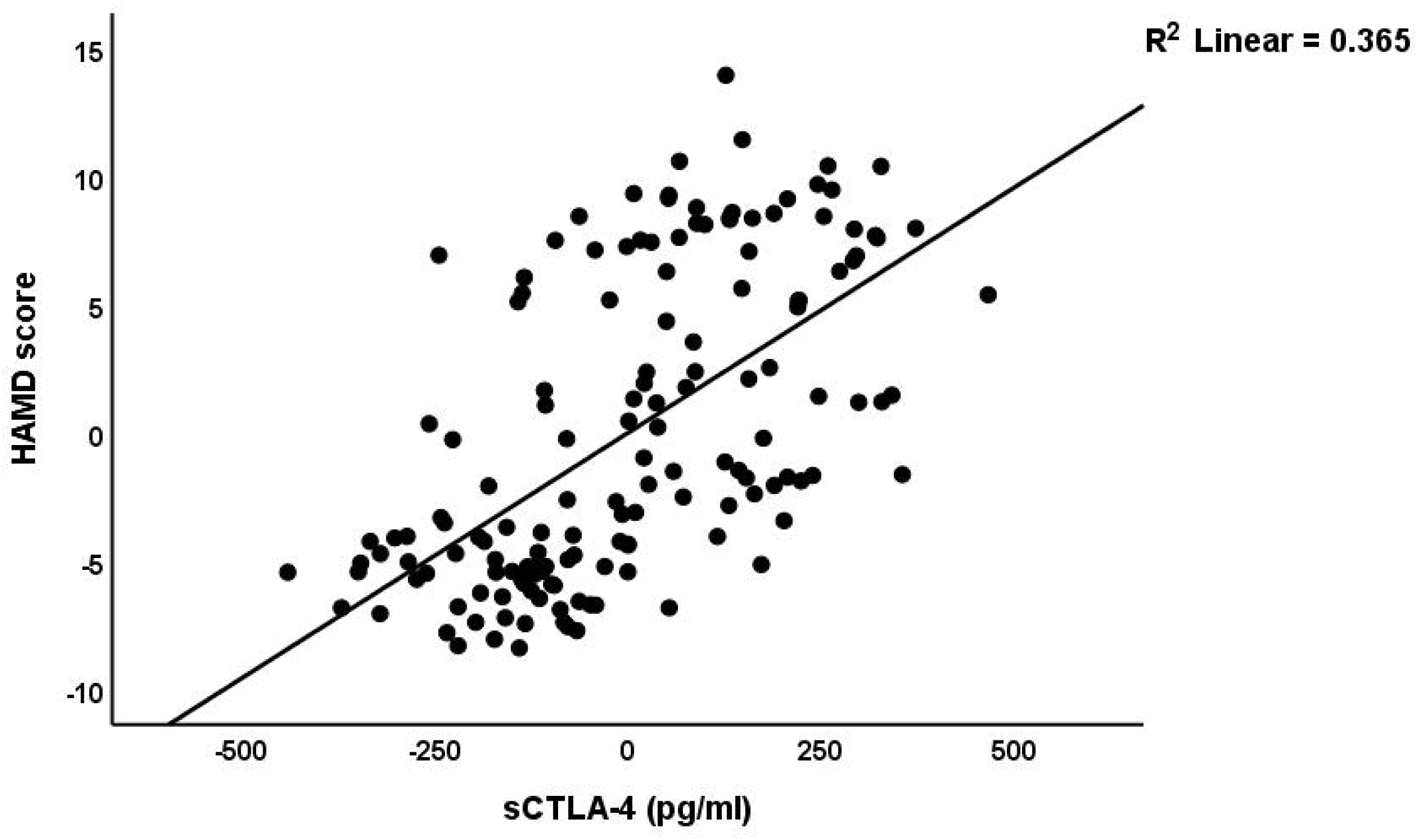
Partial regression plot of the Hamilton Depression Rating Scale (HAMD) score on serum soluble cytotoxic T-lymphocyte antigen-4 (CTLA-4) (after adjusting for age, body mass index, education) p<0.001.

**Figure 2.**
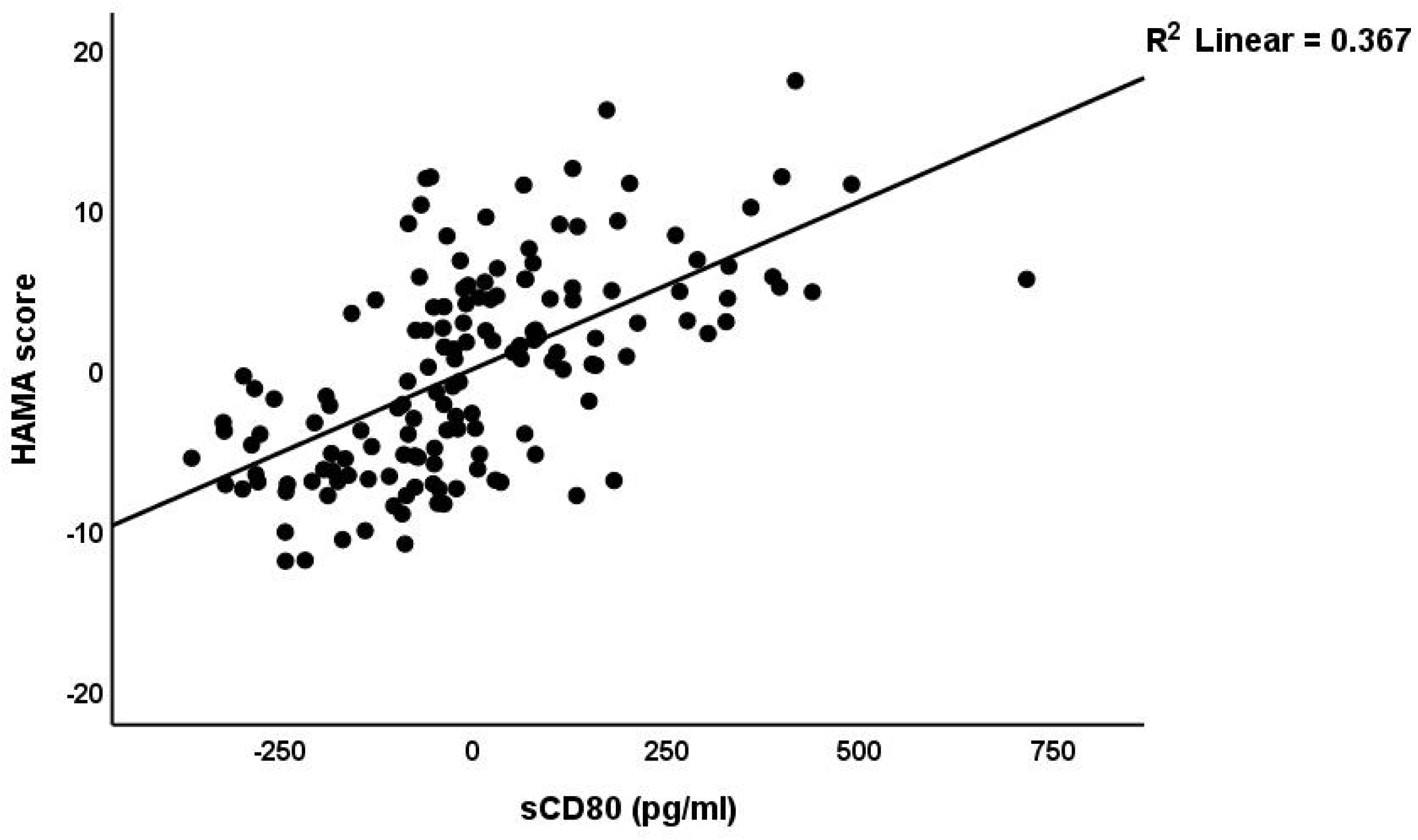
Partial regression plot of the Hamilton Anxiety Rating Scale (HAMA) score on soluble CD80 (sCD80) (after adjusting for age, body mass index, education) p<0.001.

**Table 5.** Results of multiple regression analysis with neuropsychiatric rating scale scores as dependent variables and biomarkers (without blood pressure data) as explanatory variables.

| **Regression** | **Explanatory variables** | **$\beta$** | **t** | **p** | **F<sub>model</sub>** | **df** | **p** | **R<sup>2</sup>** |
| --- | --- | --- | --- | --- | --- | --- | --- | --- |
| #1. HAMD score | **Model** |  |  |  | 32.90 | 6/143 | <0.001 | 0.580 |
|  | Vitamin D | -0.192 | -2.66 | 0.009 |  |  |  |  |
|  | sCTLA-4 | 0.360 | 5.48 | <0.001 |  |  |  |  |
|  | sCD80 | 0.267 | 3.915 | <0.001 |  |  |  |  |
|  | BMI | 0.140 | 2.51 | 0.013 |  |  |  |  |
|  | Calcium | -0.158 | -2.41 | 0.017 |  |  |  |  |
|  | GM-CSF | -0.127 | -2.30 | 0.023 |  |  |  |  |
| #2. HAMA score | **Model** |  |  |  | 36.99 | 5/144 | <0.001 | 0.562 |
|  | sCD80 | 0.345 | 5.05 | <0.001 |  |  |  |  |
|  | sCTLA-4 | 0.191 | 2.92 | 0.004 |  |  |  |  |
|  | Copper | 0.230 | 3.65 | <0.001 |  |  |  |  |
|  | Albumin | -0.178 | -2.99 | 0.003 |  |  |  |  |
|  | Vitamin D | -0.157 | -2.16 | 0.033 |  |  |  |  |
| #3. FF score | **Model** |  |  |  | 30.07 | 6/143 | <0.001 | 0.558 |
|  | Vitamin D | -0.271 | -3.64 | <0.001 |  |  |  |  |
|  | Calcium | -0.176 | -2.61 | 0.010 |  |  |  |  |
|  | sCTLA-4 | 0.202 | 3.02 | 0.003 |  |  |  |  |
|  | Copper | 0.142 | 2.27 | 0.025 |  |  |  |  |
|  | Magnesium | -0.141 | -2.44 | 0.016 |  |  |  |  |
|  | sCD80 | 0.163 | 2.29 | 0.023 |  |  |  |  |

BMI: body mass index, sCTLA-4: soluble cytotoxic T lymphocyte-associated antigen-4, GM-CSF: Granulocyte-macrophage colony-stimulating factor, HAMA: Hamilton Anxiety Rating Scale, HAMD: Hamilton Depression Rating Scale, FF: fibro fatigue scale.

The z unit based composite z sCTLA4 + z CD80 was significantly correlated with the FF (r=0.645, p<0.001), HAMA (r=0.684, p<0.001), and HAMD (r=0.702, p<0.001) scores. The sCTLA4 / sCD80 ratio was inversely and significantly correlated with the FF (r=-0.229, p=0.005), HAMA (r=-0.222, p=0.006), and HAMD (r=-0.210, p=0.010) score.

### 4. Multiple regression analyses of BP data on biomarkers

**Table 6** shows the results of the multiple regression analyses with blood pressure as dependent variable and biomarkers and clinical data as explanatory variables. Regression #1 shows that 71.6% of the variance in the systolic blood pressure can be explained by the regression on sCTLA-4, copper, and having a child (positively) and calcium and vitamin D (inversely). **Figure 3** shows the partial regression plot of the systolic blood pressure on the serum sCTLA-4. Regression #2 shows that 48.0% of the variance in the diastolic blood pressure was explained by sCTLA-4 and serum copper (both positively), and vitamin D, calcium, and GM-CSF (all negatively).

**Figure 3.**
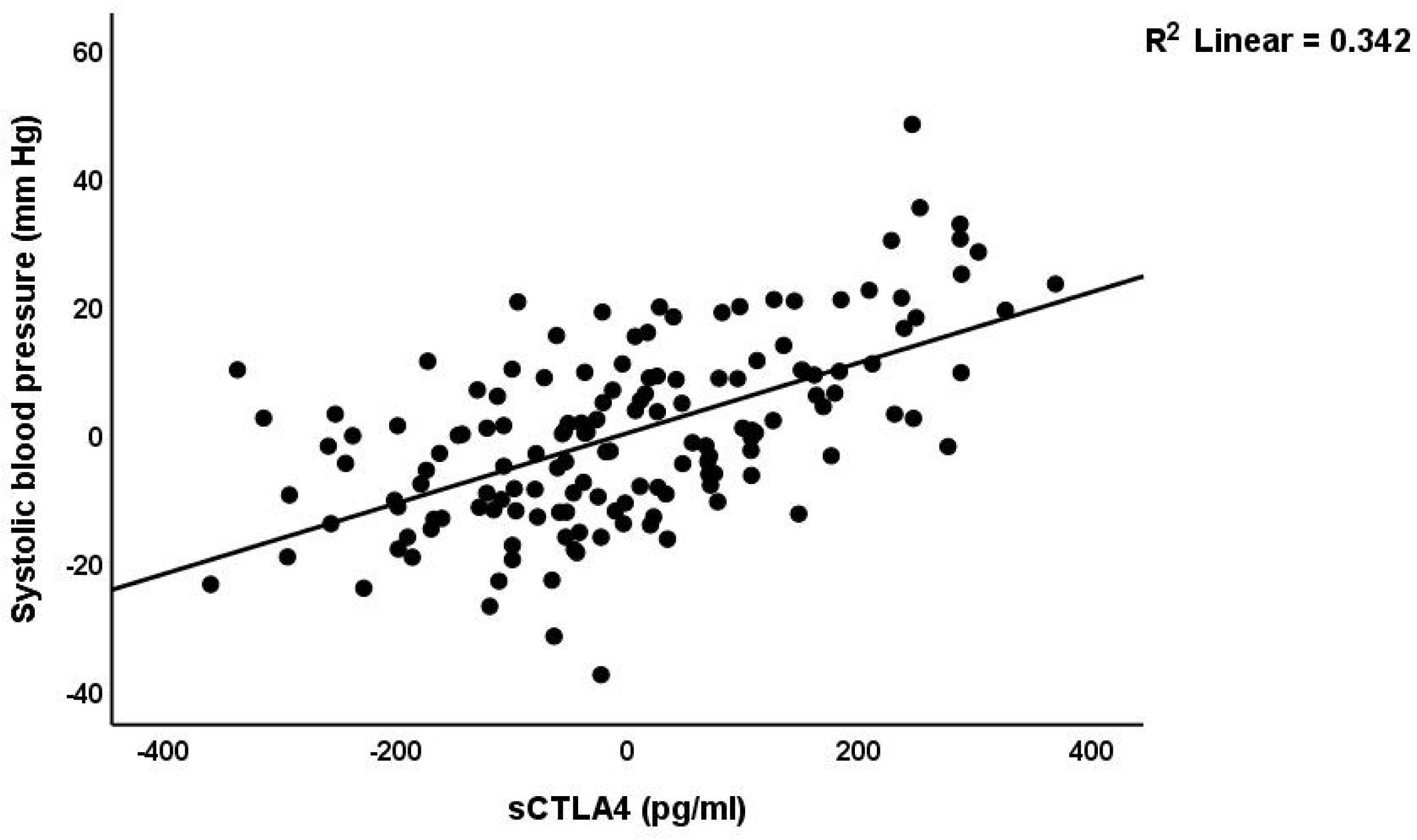
Partial regression plot of the systolic blood pressure on the serum soluble cytotoxic T-lymphocyte antigen-4 (sCTLA-4) (after adjusting for age, body mass index, calcium, vitamin D, zinc) p<0.001.

**Table 6.** Results of multiple regression analyses with blood pressure (BP) data as dependent variables and biomarkers and clinical data as explanatory variables.

| **Regression** | **Explanatory variables** | **B** | **t** | **p** | **F<sub>model</sub>** | **df** | **p** | **R<sup>2</sup>** |
| --- | --- | --- | --- | --- | --- | --- | --- | --- |
| #1. Systolic BP | **Model** |  |  |  | 75.50 | 5/144 | <0.001 | 0.716 |
|  | sCTLA-4 | 0.467 | 8.71 | <0.001 |  |  |  |  |
|  | Vitamin D | -0.266 | -4.89 | <0.001 |  |  |  |  |
|  | Calcium | -0.172 | -3.24 | 0.001 |  |  |  |  |
|  | Copper | 0.148 | 3.01 | 0.003 |  |  |  |  |
|  | Having a child | 0.109 | 2.33 | 0.021 |  |  |  |  |
| #2. Diastolic BP | **Model** |  |  |  | 26.59 | 5/144 | <0.001 | 0.480 |
|  | Vitamin D | -0.270 | -3.70 | <0.001 |  |  |  |  |
|  | sCTLA-4 | 0.276 | 3.79 | <0.001 |  |  |  |  |
|  | Copper | 0.190 | 2.86 | 0.005 |  |  |  |  |
|  | Calcium | -0.202 | -2.85 | 0.005 |  |  |  |  |
|  | GM-CSF | -0.141 | -2.31 | 0.022 |  |  |  |  |

sCTLA-4: soluble cytotoxic T lymphocyte-associated antigen-4, GM-CSF: Granulocyte-macrophage colony-stimulating factor.

## Discussion

### Increased neuropsychiatric symptoms in PE

One significant discovery from this study is that PE women exhibit higher scores in all three neuropsychiatric areas (depression, anxiety, and CFS) compared to healthy pregnant women. In this study, women with pre-existing depression, anxiety, and chronic fatigue were not included. This suggests that the findings of this research demonstrate a link between PE and the development of new neuropsychiatric symptoms.

Previous research has found that there is a significant connection between PE and the development of depression, as well as an increase in the severity of depressive symptoms (6, 60). A significant proportion of women with PE experience perinatal or postpartum depression (61). Furthermore, it has been established that PE itself is a contributing factor to the development of post-partum depression, as highlighted by the study conducted by Ye et al. (62).

Research has indicated a significant increase in anxiety levels among women who have been diagnosed with PE (63). However, a comprehensive review indicated that there seemed to be a connection between PE and depression, while no link was found with anxiety (64). Women who have experienced PE tend to report higher levels of depression and fatigue compared to those who have not had PE (10, 65).

One could make the case that the concern over potential fetal loss and other future repercussions would lead to increased levels of depression, anxiety, and fatigue in patients with PE. In addition, unexpected medical procedures and the possibility of mortality can cause feelings of depression and anxiety in pregnant women (66). However, it is worth noting that a significant portion of the variation in the severity of these neuropsychiatric symptoms can be attributed to immune-inflammatory biomarkers. This suggests that these biological factors may hold more significance than psychological factors, as will be explored in the following section.

### Biomarkers of PE

In the present study, it was observed that the PE group had higher levels of serum sCTLA-4, sCD80, and copper, while magnesium, calcium, zinc, and albumin were found to be significantly lower in PE. These findings suggest a correlation between PE and activation of immune-inflammatory responses system. PE is commonly recognized as an immune-inflammatory disorder (67, 68). Administering anti-inflammatory compounds could potentially provide benefits for women experiencing PE, as it may help address issues related to maternal immune system failure and excessive inflammation (69).

In previous studies, researchers explored the potential link between gene polymorphisms in CTLA-4 and the risk of PE. However, a meta-analysis did not yield any significant association between the two (70). In a separate study, RT-PCR revealed a decrease in the expression of checkpoint inhibitory markers, such as CTLA-4, in the decidual tissue of women with PE compared to the control group (71). Lower levels of CTLA-4 expression were observed in women with miscarriages, specifically on peripheral lymphocytes, T regulatory cells, and decidual lymphocytes. Additionally, the ratio of CTLA-4+/CD28+ in Treg cells was found to be decreased (72, 73). In patients with preeclampsia, dendritic cells exhibit elevated expression of CD80 (and CD86), which is linked to enhanced differentiation of Th1 and Th17 cells (74). Women with recurrent spontaneous abortion also show elevated levels of sCD80 (75). All in all, in the pathophysiology of PE, there is an imbalance among immune checkpoint molecules (such as CTLA-4) and stimulatory signals (74, 76, 77).

However, in this study, we focused on measuring the soluble forms of CTLA-4 and CD80 molecules, specifically sCTLA-4 and sCD80. It is important to note that these soluble forms do not per se possess the same functions as the cell-bound molecules. sCTLA-4 can suppress immune responses both in laboratory settings, animal studies, and individuals diagnosed with rheumatoid arthritis (78–81). Increased sCD80 may restore CD4+ and CD8+ T cell activation (82). Based on our findings, it seems that the decreased sCTLA-4 / sCD80 ratio in PE could suggest a shift towards heightened immune activation, possibly due to a decrease in immunosuppressive signals and a relative increase in immune-stimulatory signals. These findings align with the inflammatory biomarkers (such as decreased albumin, zinc, and magnesium) identified in our study and support the immune-inflammatory theory of PE (16–18).

Our study found no notable variation in GM-CSF levels between women with PE and those in the control group. Prior research has indicated a notable rise in serum and placental GM-CSF levels in women with preeclampsia when compared to the control group (83). In a study conducted by Gratacós et al., a similar lack of significant difference in GM-CSF was observed during the second trimester, which aligns with the findings of our own study (84). Nevertheless, at other gestational ages, there may be a significant increase in GM-CSF compared with controls (84).

It has been suggested that the decrease in calcium, magnesium, and zinc levels in the blood during pregnancy could potentially play a role in the development of PE. Therefore, adding these elements to the diet through supplementation may be beneficial in preventing PE (85). Possible reasons for the decline in these elements could be heightened inflammatory reactions and the demands of the developing fetus (86–88). One likely reason for the decrease in vitamin D levels in women who are pregnant may be the increased need for calcium metabolism to support the growth of the fetus. Several studies have consistently shown a strong link between vitamin D deficiency and a higher risk of PE (89–91), although there are some authors who do not agree with this finding (92).

### Associations between biomarkers and severity of neuropsychiatric symptoms

An essential discovery in this study is the connection observed between the neuropsychiatric rating scales and the serum biomarker levels. Therefore, a significant portion of the variation in the clinical scores (ranging from 55.8% to 58.0%) could be attributed to the presence of a combination of up to six distinct biomarkers. When it comes to predicting depression, anxiety, and chronic fatigue caused by PE, sCD80, sCTLA-4, and vitamin D are the top-3 most important predictors.

As previously mentioned, affective disorders such as major depression and associated generalized anxiety disorder, as well as CFS, are classified as neuro-immune disorders (93–97). Thus, the inverse associations between the sCTLA4 / sCD80 ratio and the FF, HAMA, and HAMD scores may play a role in the immune pathophysiology of depression, anxiety, and CFS associated with PE. It is important to emphasize that a higher T effector /T regulatory ratio plays a significant role in major depression (98). CTLA-4 is found on the surface of both T regulatory and conventional T cells, playing a crucial role in regulating the activation of T effector cells by providing negative feedback (99). In addition, CTLA-4 has the ability to compete with CD28 for ligand binding, effectively acting as a counteractive force against CD28-mediated co-stimulation (100). Interestingly, some, but not all studies reported a significant association between CTLA-4 gene polymorphisms and major depression (101, 102). Decreased levels of zinc, magnesium, and vitamin D, along with elevated copper, are significant indicators of conditions such as depression, perinatal depression, anxiety, CFS, and perinatal fatigue (103–113).

One noteworthy discovery from the present study is the strong predictive power of sCTLA-4, copper, calcium, and vitamin D in relation to systolic and diastolic hypertension. Previously, it was shown that there is a connection between depressive symptoms in early pregnancy and the mother’s blood pressure during the first trimester (9). Other researchers have found that women with higher blood pressure in the third trimester tend to experience increased depression (5, 62). There is a correlation between immune activation and oxidative and nitrosative stress, which has been linked to hypertension in individuals with depression (114). Inflammation is linked to hypertension through the activation of pathways related to oxidative stress, immune activation caused by sodium, and the inflammasome (115). Vitamin D plays a crucial role in promoting angiogenesis and reducing blood pressure by affecting the renin-angiotensin system (91, 116–120). Therefore, vitamin D is expected to play a role in repairing the endothelium and promoting angiogenesis, while also regulating blood pressure (121).

## Limitations of the study

It would have been intriguing to evaluate T effector and T regulatory cells through flow cytometry, as well as assess membrane-bound CTLA-4, CD28, CD80, and CD86 on T cells. It would be beneficial to conduct further analyses on oxidative and nitrosative stress. One could make the case that the sample size is relatively small. Nevertheless, the sample size was determined through power analysis, and the subsequent power achieved in the primary outcome variables (as analyzed through multiple regression on the biomarkers, as shown in Table 5) was 1.0.

## Conclusions

Compared to control women, PE women exhibit higher depression, anxiety, and CFS scores. Approximately 55.8%-58.0% of the variance in the HAMD, HAMA, and FF scores was accounted for by the regression on biomarkers, and sCTLA-4, sCD80, and vitamin D were the three most significant biomarkers. The HAMD/HAMA/FF scores exhibited a significant and inverse correlation with the sCTLA-4/sCD80 ratio. Approximately 70% of the variance in systolic blood pressure was predicted by copper, sCTLA-4, vitamin D, and calcium. The results emphasize that symptoms of depression, anxiety, and chronic fatigue associated with PE are accompanied by immune-inflammatory response activation. Imbalances among soluble checkpoint molecules contribute to the pathogenesis of both hypertension and neuropsychiatric symptoms associated with PE. sCTLA-4 and membrane CTLA-4 as well as sCD80 and membrane CD80 are new drug targets to treat PE and depression, anxiety, and CFS due to PE. Moreover, sCTLA-4 and copper, and lowered calcium and vitamin D are new drug targets to treat hypertension in PE women.

## Acknowledgments

We thank the staff of the Dialysis Unit at Al-Hakeem General Hospital for helping in collecting samples and the Asia Clinical Laboratory in Najaf for hematological and biochemical testing.

## Author’s contributions

All the contributing authors have participated in the preparation of the manuscript.

## Data availability statement

The database created during this investigation will be provided by the corresponding author (MM) upon a reasonable request once the authors have thoroughly used the data set.

